# High Fidelity, CT on Rails-based Characterization of Total Delivered Dose Variation for Conformal Head and Neck Treatment: With evaluation of adaptive replanning time-point implications

**DOI:** 10.1101/2023.04.07.23288305

**Authors:** Haocheng Dai, Vikren Sarkar, Christian Dial, Markus D. Foote, Sarang Joshi, Bill J. Salter

**Affiliations:** Scientific Computing and Imaging Institute, University of Utah, Salt Lake City, UT 84112, U.S.A.; Department of Radiation Oncology, University of Utah, Salt Lake City, UT 84112, U.S.A.

## Abstract

**Purpose:** This study aims to characterize dose variations from the original plan for a cohort of head-and-neck cancer (HNC) patients using high-quality computed tomography on rails (CTOR) datasets and evaluate a predictive model for identifying patients needing re-planning.

**Material and methods:** 74 HNC patients treated on our CTOR equipped machine were evaluated in this retrospective study. Patients were treated at our facility using in-room, CTOR Image Guidance — acquiring CTOR kV fan beam CT (FBCT) images on a weekly to near-daily basis. For each patient, a particular day’s delivered treatment dose was calculated by applying the approved, planned beam set to the post image-guided alignment CT image-of-the-day. Total accumulated delivered dose distributions were calculated and compared to the planned dose distribution and differences were characterized by comparison of dose and biological response statistics.

**Results:** The majority of patients in the study saw excellent agreement between planned and delivered dose distribution in targets — the mean deviations of D95 and D98 of the planning target volumes (PTVs) of the cohort are −0.7% and −1.3%, respectively. In critical organs, we saw a +6.5% mean deviation of mean dose in parotid glands, −2.3% mean deviation of maximum dose in brainstem, and +0.7% mean deviation of maximum dose in spinal cord. 10 of 74 patients experienced nontrivial variation of delivered parotid dose which resulted in a normal tissue complication probability (NTCP) increase compared to the anticipated NTCP in the original plan, ranging from 11% to 44%.

**Conclusion:** We determined that a mid-course evaluation of dose deviation was not effective in predicting the need of re-planning for our patient cohorts. The observed non-trivial dose difference to parotid gland delivered dose suggest that even when rigorous, high quality image guidance is performed, clinically concerning variations to predicted dose delivery can still occur.

## 1. Introduction

Conformal radiation therapy is a highly effective treatment approach for many cancers. Intensity-modulated radiation therapy (IMRT) allows for a more precise conformation of radiation dose to the targeted tumor volume and increased sparing of surrounding normal tissues (Chao et al. 2001; Eisbruch et al. 2010; N. Lee et al. 2002). Due to high sensitivity of head and neck tissues, and subsequent potential for non-trivial side effects, it is imperative that the high dose region be delivered with high accuracy and consistency (Barnett et al. 2009). Advanced image-guided radiotherapy (IGRT) techniques, such as cone beam computed tomography (CBCT) and in-room CT on rails (CTOR) increase the accuracy of dose delivery, thereby helping to ensure the fidelity of the delivered dose distribution relative to the planned distribution. However, even the best IGRT approach cannot undo anatomical changes, such as weight loss or tumor shrinkage, that occur in patients as the radiation course progresses. These changes can compromise target coverage or increase dose to sensitive structures.

Although dose deviations from planned distributions are known to occur throughout the treatment and have been previously studied (O’Daniel et al. 2007), the accuracy of such evaluations are inherently limited by the quality of the in-room, daily imaging modality. Even when 3D imaging is obtained daily via CBCT, the reduced spatial, contrast and Hounsfield-unit (HU) resolution of CBCT, relative to the fan beam CT (FBCT) simulation dataset, limits the precision with which dose variation can be studied. In particular, the increased scatter component of CBCT imaging influences the relationship between HU, attenuation coefficient, and electron density of patient tissues (Giacometti et al. 2019) resulting in increased uncertainty in dose calculations when compared with FBCT images.

In previous studies (C. Lee et al. 2008; Barker et al. 2004; Beltran et al. 2012; Hunter et al. 2013) on dose tracking, a limited number of patients (ranging from 10 to 18) were included in the study cohorts which used in-room tomotherapy megavoltage CT (MVCT) (C. Lee et al. 2008), integrated CT-linear accelerator system (EXaCT) (Barker et al. 2004), return of patient to the CT simulator, kV CBCT (Hunter et al. 2013) to image anatomical changes during the treatment course. To address the data deficiency problem and to get a better understanding of potential dose deviation, McCulloch et al. (McCulloch et al. 2019) built a larger cohort of 100 patients. This added clinical variety and significance to the evaluation, however, the daily imaging modality was still limited to kV CBCT and full 3D dose accumulation was not available, necessitating an approximation to estimate the accumulated dose and thus introducing additional uncertainties into the evaluation.

In an effort to improve upon the accuracy and reliability of recalculated dose distributions, we present accumulated, full-course dose distributions for 74 HNC patients treated at our facility using gold-standard in-room, fan beam CTOR Image Guidance. Acquisition of CTOR kV FBCT images on a weekly to near-daily basis for these patients has enabled us to compile a large, gold-standard FBCT dataset for dose variation investigation. In addition to improved dose-calculation accuracy, the FBCT-to-FBCT image registration employed here, facilitates more accurate structure mapping between CT-of-the-day and simulation planning CT. In combination with a FBCT-based, high fidelity, full dose recalculation, this allows for improved accuracy in dose tracking and summation that is comparable in accuracy to that of the original plan.

Moreover, such high-accuracy reconstruction of delivered dose ensures improved accuracy in the characterization of dose variations from the original plan. This improved understanding of delivered dose variation, in turn, facilitates improved insights into circumstances leading to observed side effects, along with an evolved rationale for adaptive re-planning time points.

## 2. Material and Methods

### 2.1. Patient Data

Our novel dataset consists of 74 patients with HNC treated between 2012 and 2020. The study was approved by the institutional review board. For each patient, there is one planning FBCT simulation scan, one approved and delivered treatment plan, and 10 − 39 daily FBCT IGRT image sets, with an average of 31.1 daily FBCTs per patient. The frequency of imaging in the patient cohort ranges from 1 to 2.7 days, with more than half of the patients receiving a FBCT at least every second day during the treatment.

All patients were originally planned in the Eclipse treatment planning system (TPS) (version 11.0.42, Varian Medical Systems, Palo Alto, CA) and dose was calculated using the anisotropic analytical algorithm (AAA). Additional plan details and patient demographics are detailed in Table 1.

**Table 1.** Demographic of our patient cohort

| **Patient cohort** |  |
| --- | --- |
| Patients, $n$ | 74 |
| Sex, $n$ | |
| Male | 63 (85.1%) |
| Female | 11 (14.9%) |
| Age, $y$ | |
| <30 | 1 (1.4%) |
| 30-39 | 3 (4.0%) |
| 40-49 | 6 (8.1%) |
| 50-59 | 19 (25.7%) |
| 60-69 | 37 (50.0%) |
| 70-79 | 8 (10.8%) |
| Fractions, $n$ | |
| 30 | 49 (66.2%) |
| 33 | 9 (12.2%) |
| 35 | 9 (12.2%) |
| 39 | 4 (5.4%) |
| Others | 3 (4.0%) |
| Prescribed dose, cGy |  |
| 6000 | 8 (10.8%) |
| 6600 | 23 (31.0%) |
| 6750 | 29 (39.2%) |
| 7000 | 9 (12.2%) |
| 7020 | 4 (5.4%) |
| Others | 1 (1.4%) |

### 2.2. Image guided radiotherapy

Patients were treated on a Siemens Artiste linear accelerator equipped with an inroom Siemens CTOR scanner (SOMATOM Sensation 40, Siemens Healthcare, Erlangen, Germany) which was used for pretreatment imaging and positioning.

### 2.3. Dose tracking workflow

Dose tracking was carried out using the RayStation TPS (Bodensteiner 2018) (version 10A, RaySearch Laboratories AB, Stockholm, Sweden) and was automated using the built-in scripting application programing interface (API). Original planned dose distributions were recalculated in RayStation prior to starting dose accumulation. Several scripts were developed to automate the following steps: replicate the registration utilized for image guidance, deformable image registration (DIR), contour propagation, dose calculation on daily images, dose deformation, and dose accumulation.

Rigid registrations utilized for daily image guidance are stored in digital imaging and communications in medicine (DICOM) files as a frame-of-reference transformation matrix and include the operations of translation and rotation. Registrations were loaded into RayStation, along with daily images, then applied to reproduce daily setup, and map beams to CTs-of-the-day for daily dose calculation.

DIR was carried out using a hybrid deformable registration technique (ANACONDA) (Weistrand and Svensson 2014) in RayStation which combines image intensity and anatomical information (including ROIs and points of interest (POIs) together). In our implementation, the anatomical information was not used in the registration technique and the objective function consisted only of image similarity and grid regularization terms.

After calculating the deformation map, the OAR and target contours were propagated to the daily CT space using the deformation field. Dose was then calculated using the RayStation collapsed cone algorithm (Ahnesjö 1989) on each daily CT image to estimate the actual delivered dose distribution for each treatment session. Subsequently, daily doses were deformed back to the planning CT and accumulated to allow for direct comparison against the planned dose distribution. For treatment days without daily CT images, the most recent prior dose calculation was repeated in the accumulation.

To circumvent potential errors in dose evaluation related to variations of field of view (FOV) in daily images, a sequence of contours that delineated the FOVs in both the planning CT and daily CTs were generated. Deformation mappings computed earlier were then leveraged to map the FOV contours from the daily CT’s space to the planning CT space. After all the FOV contours were presented in the same reference space, the intersection was calculated and rendered as the common FOV contour. The intersection of individual target and OAR structures with the common FOV were subsequently calculated to ensure that daily dose volumes encompassed relevant structures.

### 2.4. Image registration validation

The image registration procedure in RayStation consists of two parts: rigid registration and deformable registration. To verify if the rigid registration was performing well, we reviewed all planning CT and daily CT pairs and confirmed that the ANACONDA algorithm was performing well in all of the 74 patients’ data — i.e. the bony structures were well aligned without any visible misalignment. For the deformable registration, we visually verified the resulting deformed daily CT across the dataset and confirmed that the algorithm was manifesting robustness, even when handling large but reasonable anatomical changes. Both of these reviews were performed by a senior medical physicist with extensive expertise in image guidance and registration. We note that the ANACONDA algorithm was previously validated by Weistrand et al. (Weistrand and Svensson 2014) on CBCT data of the head and neck region and was reported to have performed well in comparison with other algorithms in DIR-LAB.

### 2.5. Dosimetric evaluation

Dose that was accumulated onto the original simulation planning FBCT was used for all characterizations of the summed, delivered dose. Target coverage was evaluated in terms of dose received by 95% (D95%) and 98% (D98%) of the volume. OAR evaluations include mean dose to parotid glands, maximum dose to brainstem, and maximum dose to spinal cord. Relative deviations for all metrics are reported as below:

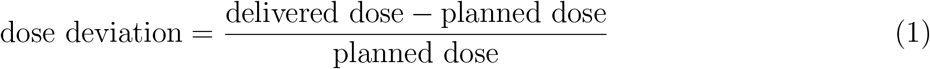

### 2.6. Biological response

With an aim to identify the cases where the parotid gland would experience a high (and subsequently increased) probability of complication due to observed increase in delivered dose, we calculated normal tissue complication probability (NTCP) for a subset of patients that exceeded planning criteria. Specifically, we filtered out patients with parotid glands where the original planned mean dose was larger than 26 Gy (our planning goal), intersected with those patients where the difference between delivered and planned mean dose was also increased by 4 Gy or more (suggested overdose threshold for re-planning by Hunter et. al. (Hunter et al. 2013)), which was intended to yield insight into the biological manifestations of variations in delivered dose.

Here, we used the RayStation NTCP-Poisson LQ models (Källman, Ågren, and Brahme 1992; Brahme 1984; Levin-Plotnik et al. 2001; Lind and Mavroidis 1999) for NTCP evaluation (xerostomia endpoint). For parotid glands’ NTCP metric, we set the maximum normalized gradient of the dose response curve at 1.8 and the dose giving a 50% response probability D50 at 46 Gy (Källman, Ågren, and Brahme 1992). As the NTCP of planned and delivered doses were both evaluated in the same simulation planning CT space, the uncertainty of the biological response deviation stems only from the dose received by each voxel at each fraction.

## 3. Results

Deviations of D95/D98 for the 190 planning target volumes (PTV) are shown in Figure 1. Note that due to some patients being treated bi-laterally or with simultaneous integrated boosts, one patient may have more than one PTV contour. The mean deviations of D95 and D98 of the PTVs were observed to be −0.7% and −1.3%, respectively. Among patients whose PTVs experienced decreased D95, the maximum deviation was −12.0%, followed by a patient whose D95 variation was −8.7%. With regard to D98 evaluation, 12 patients’ PTVs experienced greater than 10% decrease, the largest followed by the next largest deviations of −28.3% and −16.8%, respectively. We closely investigated the patients whose PTVs experienced more than a 10% decrease in D98 and listed them in Table 2. As is shown, patients who saw more than a 10% decrease in D98 of PTVs did not observe the same degree of deviation in D95 due to the relative dose shift being limited to within 10%. In contrast, the PTV of patient HN013 experienced a 12% decrease in D95 but saw a 13.7% increase in D98.

**Figure 1.**
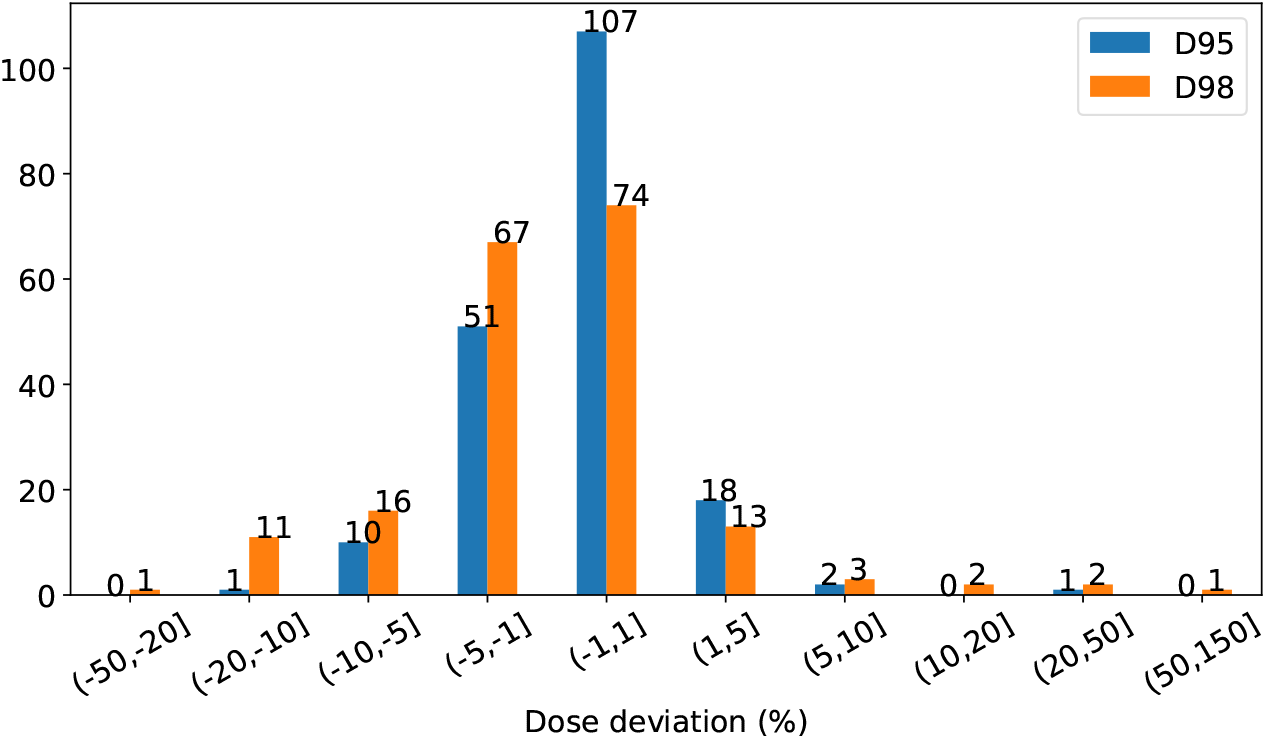
Distribution of D95 and D98 deviation in PTVs. A positive percentage indicates an increase from the plan and a negative percentage depicts a decrease from the plan.

**Table 2.**
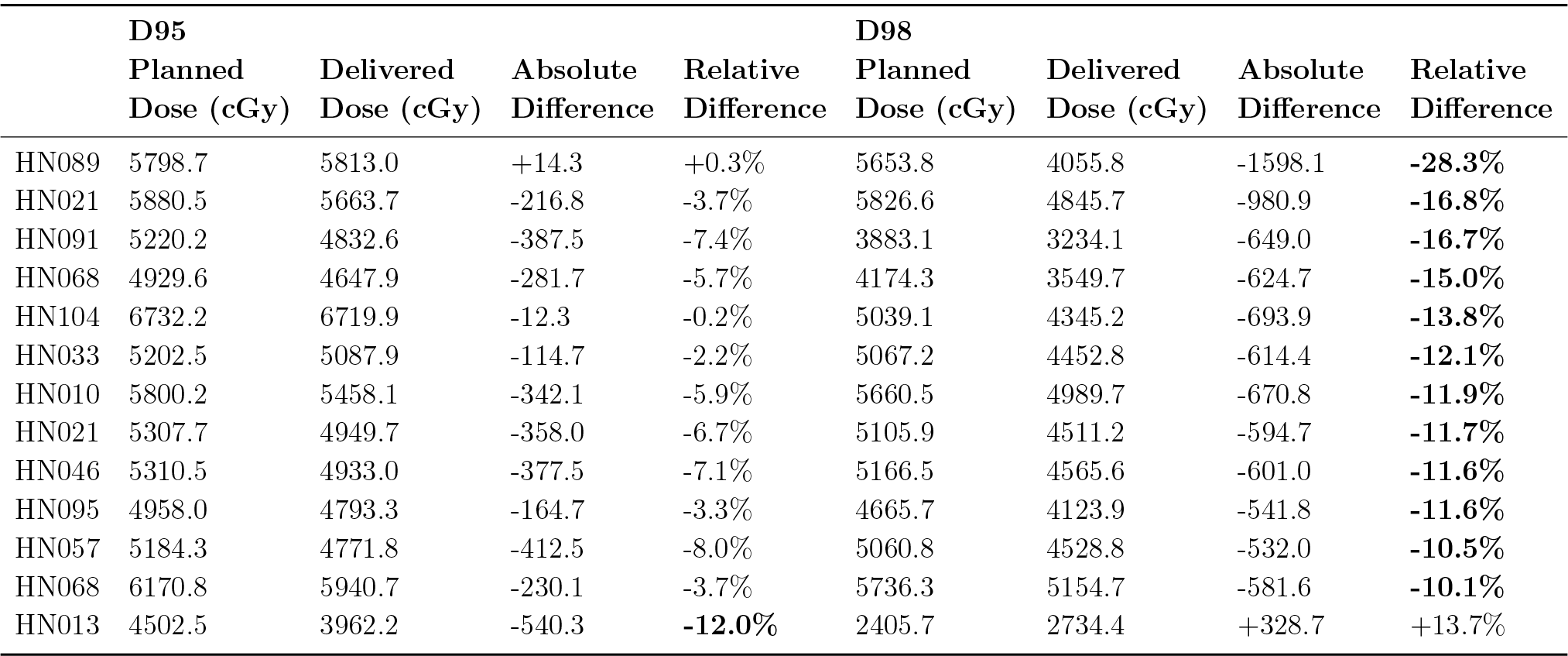
Patients with PTVs where D95 or D98 decreased by more than 10% (bold font). A positive percentage indicates an increase from the plan and a negative percentage depicts a decrease from the plan.

Total delivered dose was evaluated for 147 parotid glands (one patient had only a right parotid gland), 73 brainstems (one patient’s brainstem contour was not transferred), and 74 spinal cords. The distribution of variation between planned and delivered dose of critical OARs is detailed in Figure 2.

**Figure 2.**
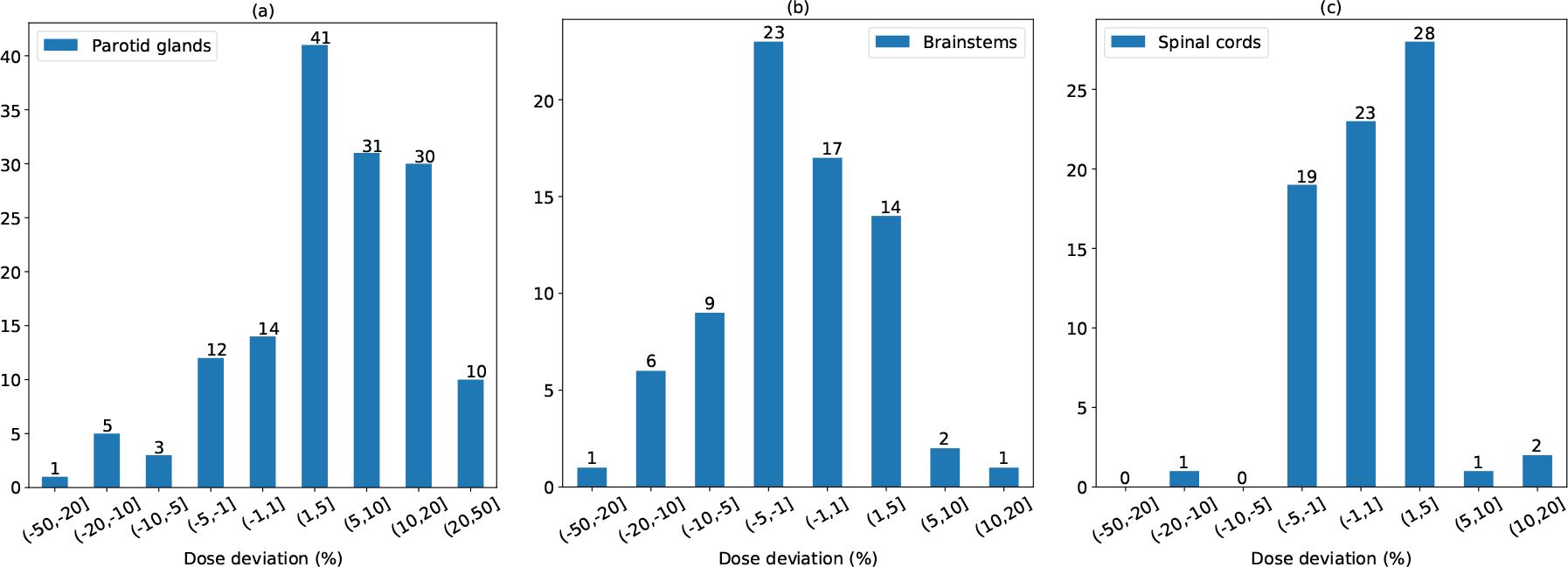
(a): distribution of mean dose deviation in parotid glands; (b): distribution of maximum dose deviation in brainstems; (c): distribution of maximum dose deviation in spinal cords. A positive percentage indicates an increase from the plan and a negative percentage depicts a decrease from the plan.

We observed an average 6.5% increase of mean dose across all 147 parotid glands for all 74 patients. Seventy one of the 147 parotid glands experienced a 5% or greater increase in mean dose, with 10 (13.5%) receiving 20% − 50% higher mean dose than that was indicated by the original treatment plan. Less frequent increased dosing of the brainstem was observed: the mean and maximum deviation of maximum dose was a 2.3% decrease, and a 12.5% increase, respectively. Only two patients experienced a 5 − 10% increase in maximum dose to the brainstem, with one receiving a cumulated maximum dose that was 12.5% higher than indicated by the original treatment plan. We note that, while the dose to brainstem increased above what was originally planned, it is well below the known tolerance dose for this structure. For the spinal cord, the mean and maximum deviation of maximum dose was a 0.7% increase and a 13.7% increase, with 96% of patients receiving less than 5% relative increase above the originally planned maximum delivered dose. Again, an increase in delivered dose beyond what was originally predicted, does not mean that the structure exceeded its known tolerance dose.

As demonstrated in the Figure 2(b)(c), for maximum dose, only 3 patients’ brainstems and 3 patients’ spinal cords received 5% or more dose than was originally planned and approved. The majority of total dose increases occurred in the parotid glands, as priority is typically given to adequate dose coverage of the target, which can subsequently and (sometimes) unavoidably spill dose to the immediately adjacent parotid gland(s).

We next curated a set of patients of interest who had at least one parotid gland that was prescribed greater than 26 Gy mean, initial planning dose, and for which the subsequent delivered mean dose was even higher than the planning goal by more than 4 Gy (Hunter et al. 2013). The dosimetric difference for parotid glands in this patient subset is listed in Table 3. Ten of 74 patients experienced a nontrivial variation of the delivered dose in parotid glands (according to the previously stated criteria), which resulted in NTCP increases compared to the anticipated NTCP in the original plan, ranging from 11% to 44%. Table 3 lists the planned and delivered NTCPs in parotid glands. Notably, the NTCP in the right parotid glands increased 44% and 27% in HN010 and HN030, and the NTCP of HN010 in the left parotid glands increased 23%, which suggests a potential, negative biological response in patients’ bodies.

**Table 3.**
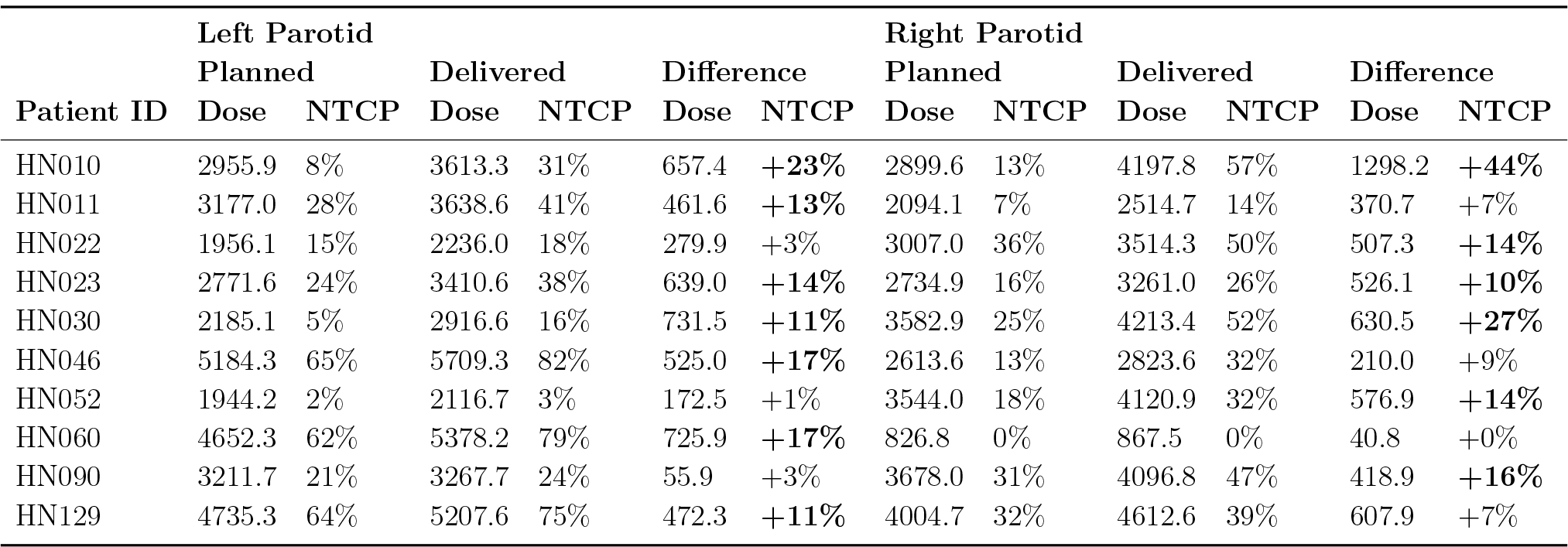
Comparison of mean dose (cGy) and NTCP of parotid glands in planned and delivered doses. Bold font suggests a more than 10% NTCP increase. A positive percentage indicates an increase from the plan and a negative percentage depicts a decrease from the plan.

## 4. Discussion

In this study, we used in-room, CTOR-generated, fan-beam-CT data sets, equivalent to the high fidelity, fan beam simulation-CT data sets used for original plan calculation to recalculate daily variations to the dose actually delivered to 74 HNC patients during over 2,200 treatment fractions, using an average of 31 daily imaging sessions per patient. The use of gold standard fan beam data sets from in-room CTOR ensures that the daily dose variations we characterized are equivalent in fidelity to the original, planned dose distributions, overcoming the limitations of other approaches that used less accurate CBCT-generated dose recalculations.

The presented data confirms that the vast majority of patients treated using high-quality daily image guidance receive delivered dose distributions that are very consistent with the originally planned treatment. However, non-trivial variations in delivered doses were still observed for multiple patients. Non-trivial increases to the parotid gland demonstrate that even when rigorous, high-quality image guidance is performed, clinically concerning variations to predicted dose delivery can occur.

With regard to dose variation, significantly more patients experienced increases in delivered dose (versus decreased dose) of the parotid gland, which is reasonable when we consider that typical planning isodose distributions achieve full coverage of the immediately adjacent target area by carefully carving out a narrow window of sparing for the parotid gland. Any variation or change in patient body habitus (e.g. weight loss) can easily cause the previously protected parotid gland to shift into the high dose region, and thus be overdosed.

While significant increases of delivered dose, relative to planned dose, are of obvious potential concern, the most important factor to consider is the biological impact. We curated 10 patients (13.5%) as patients of interest, in order to better characterize biological impact, for which we subsequently calculated the NTCP for a xerostomia endpoint. Remarkably, two of the 10 patients of interest experienced greater than 25% increase in the original probability of xerostomia (27% and 44%, respectively), which characterizes the clinically significant increase in risk to the patient versus simple quantification of delivered dose variation.

In order to evaluate the potential need to re-plan during the treatment course, we also explored the correlation between the dosimetric data at the middle and the end of the treatment, as previously proposed by Hunter et. al. (Hunter et al. 2013) and McCulloch et. al. (McCulloch et al. 2019), where it was suggested that a mid-course dose deviation is likely to be predictive of the outcome for the entire treatment course. Recently published data from McCulloch et. al. (ibid.) suggested that a < 15% deviation between the planned and delivered dose for parotid glands would not have a significant toxicity impact in a patient population. While this threshold may be debatable, we endeavored to investigate the validity of this assertion for our own dataset. In Figure 3, we plot the deviation from the total planned to total delivered mean dose received by 147 parotid glands of 74 patients. Additionally, we calculated the deviation from accumulated planned dose to accumulated delivered mean dose in the first half of the treatment for each parotid gland and scaled each by 2 to serve as the predicted deviation at the end of the treatment (magenta dots in Figure 3). The predicted mean dose deviation at the end of the treatment is connected by color-coded segments to the actual observed total mean dose deviation at the end of the treatment. We ordered the parotid gland data by the predicted mean dose deviation for easy illustration and understanding. As can be seen, 10 parotid glands’ predicted mean dose deviation exceeded a 3.9 Gy dose deviation threshold, i.e. 15% of the 26 Gy prescribed mean dose for parotid glands. Among these, three parotid glands saw a decrease from predicted deviation to observed total dose deviation, and for one the observed total mean dose deviation fell under the 3.9 Gy threshold suggesting a 90% positive predictive value (PPV) in the cohort. For the parotid glands where the predicted dose deviation did not surpass the 3.9 Gy threshold, 14 of these saw the observed final dose deviation reach above the threshold, indicating an 89.8% negative predictive value (NPV). Thus, the sensitivity of this model is only 43.5%, despite a 99.2% specificity.

**Figure 3.**
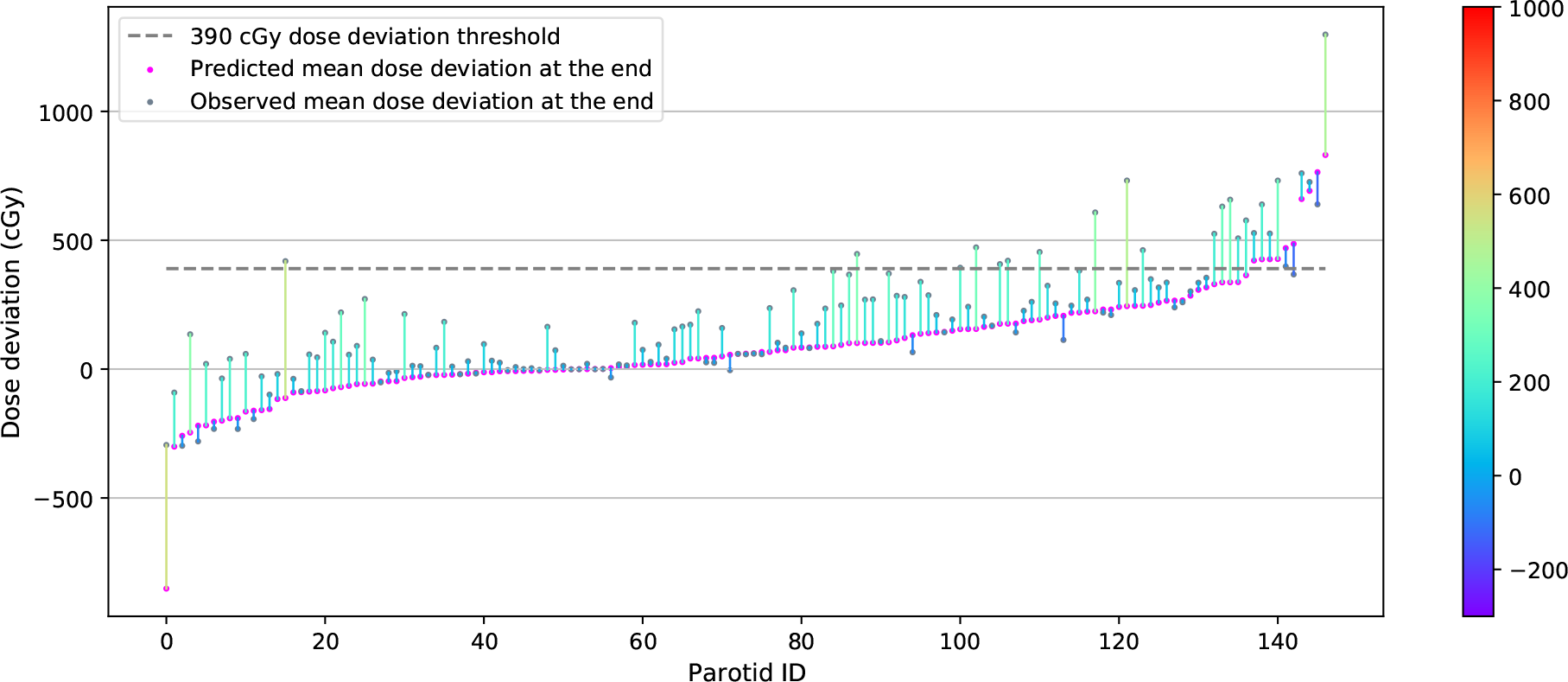
Correlation between predicted mean dose deviation and observed total mean dose deviation. Parotid glands are sorted ascendingly from left to right by predicted mean dose deviation at the end of the treatment (magenta points). The corresponding observed total mean dose deviations (gray points) are connected by color-coded segments to predicted mean dose. The color bar on right indicates the magnitude of discrepancy between the predicted value and observed outcome.

In an effort to ascertain a proper threshold for predicting the need of re-planning, we reorganized the parotid gland data in Figure 3 and visualized them in Figure 4 by sorting the parotid gland ascendingly from left to right by observed total mean dose deviation at the end of the treatment. We endeavored to investigate the pattern of the corresponding mid-course dose deviation of the parotid glands whose final deviation is above 3.9 Gy (mid-course deviation threshold at 1.95 Gy). The minimum dose deviation of this group of parotid glands at the midpoint of the treatment was −56 cGy, which indicates that after going through the first half of the treatment, the actual delivered mean dose was even lower than the prescribed mean dose. In Figure 4, in spite of the upward trend of the gray dots representing different parotid glands’ observed total mean dose deviation, we could not observe an upward trend in mid-course dose deviation (magenta points), regardless of the variance of the mid-course deviation increases with the trend of total dose deviation.

**Figure 4.**
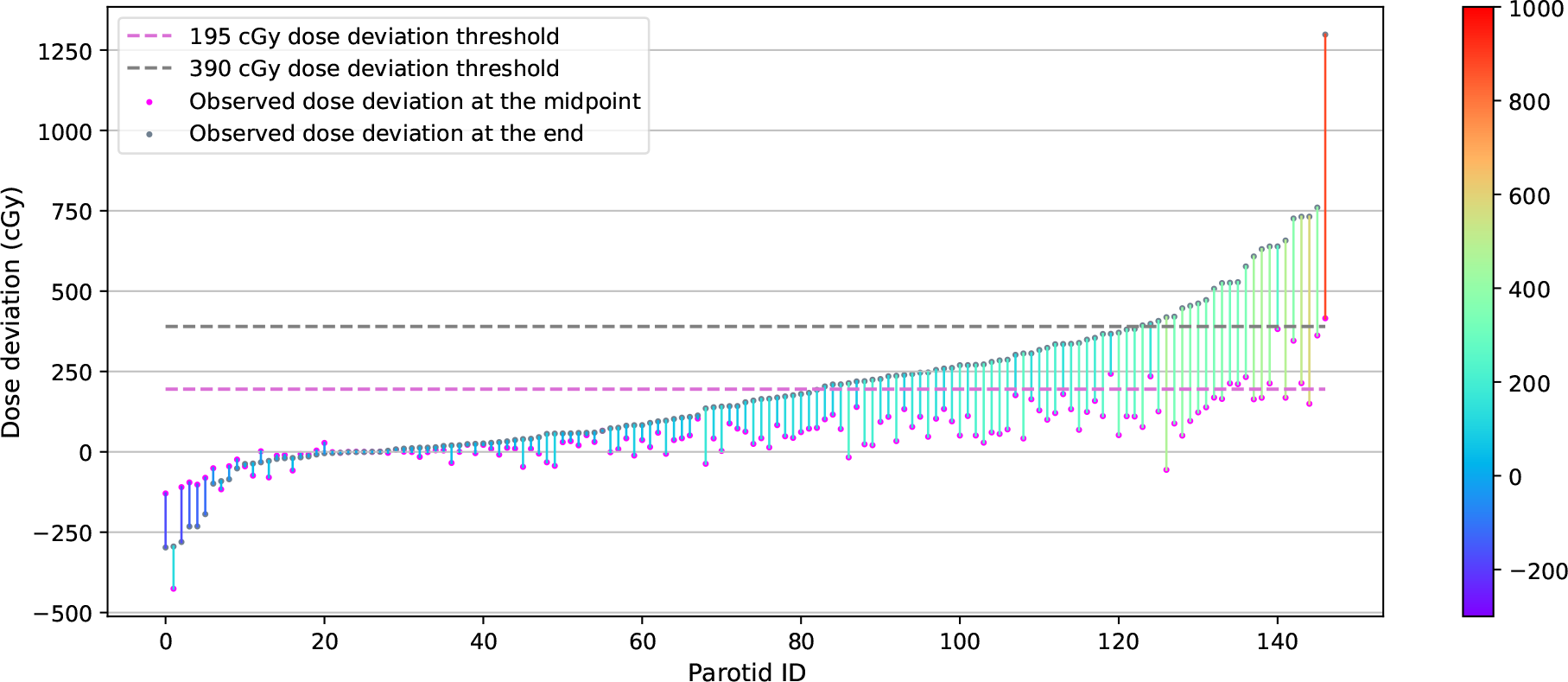
Correlation between observed mean dose deviation at midpoint of the treatment and at the end of the treatment. Parotid glands are sorted ascendingly from left to right by observed mean dose deviation at the end of the treatment (gray points). The corresponding mid-course mean dose deviations (magenta points) are connected by color-coded segments to total mean dose deviation. The color bar on right indicates the magnitude of discrepancy between the predicted value and observed outcome.

In the interest of identifying a specific threshold model to identify patients in need of re-planning, we also investigated the correlation between predicted and actual total dose deviation. In Figure 5(a), we plot 147 parotid glands in the 2D space with predicted total dose deviation on the *x*-axis and actual total dose deviation on the *y*-axis. The color of the sample points read as

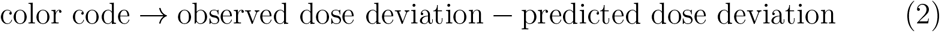

The more red the data points are, the larger the positive dose difference that exists. The more blue the data points are, the larger the negative dose difference that exists. We leveraged a linear regression model to represent the correlation of the two variables and visualized it as the yellow dashed line. To yield a more intuitive illustration, we also plot a green dashed line with slope = 1 and *y*-intercept = 0. From Figure 5(a) we observe that one hundred and sixteen parotid glands saw an increase from predicted total dose deviation to observed total dose deviation (points located above the green dashed line) and 72 out of 147 parotid glands had a positive predicted dose deviation and an even larger actual dose deviation. We observed that the majority of parotid glands (78.9%) received more dose in the latter half of the treatment than in the first half of the treatment. This observed pattern is consistent with our experience: many patients may experience weight loss resulting from the first half of the treatment, which can lead to a larger variation from the initial calculated dose in the latter half of the treatment.

**Figure 5.**
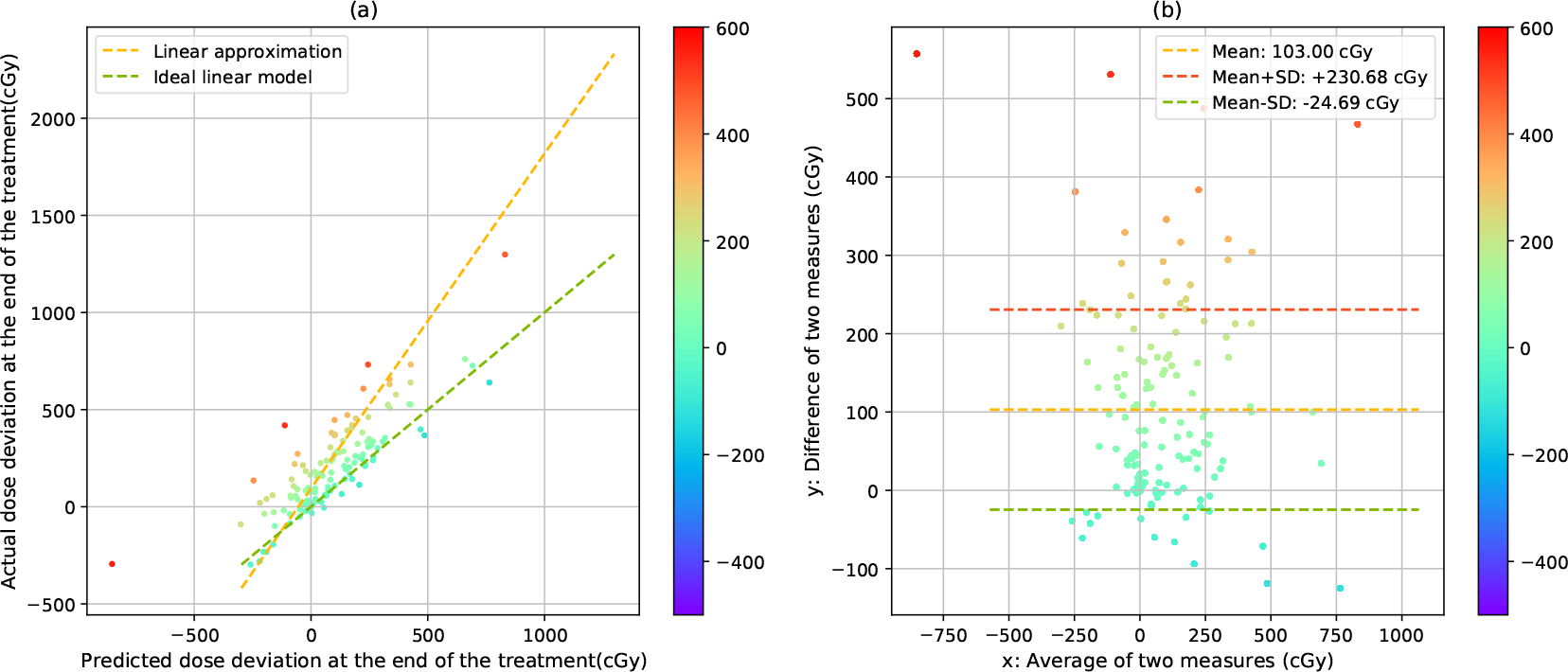
(a): 2D distribution of parotid glands in the space of final actual dose deviation-final predicted dose deviation at the end. Points are color-coded by the residual to the ideal linear model. (b): Bland-Altman plot of actual dose deviation and predicted dose deviation in parotid glands. Points are color-coded by their distance to 0 difference of two measures lines.

In Figure 5(b), we present a Bland-Altman plot to analyze the agreement between the observed dose deviation and predicted dose deviation. Ideally, a gold standard measure should have all the sample points located on the *y* = 0 line (difference of two measures equals to 0). However, as shown in Figure 5(b), most of the sample points are located above the *y* = 0 line, with the mean difference equal to 103.0 cGy and a standard deviation of 127.7 cGy. Therefore, we do not find any special pattern of the distribution w.r.t. the average of the two measures, i.e. the data points are rather evenly distributed along the average of two measures’ axes. Or, in other words, the difference between the predicted and observed total dose deviation is not strongly correlated with the predicted total dose deviation; therefore, a mid-course evaluation of the need for re-planning is unable to predict overdosing of critical OARs at the end of the treatment.

The high fidelity, CTOR-derived, delivered dose data presented here makes clear that a limited subset of patients may experience clinically relevant increases in delivered dose, even when these patients are treated with daily, high-resolution image guidance. Our data further confirms that, through the use of gold standard FBCT-based dose recalculation, some degree of adaptive re-planning will be needed for a subset of patients and, furthermore, a mid-course evaluation of dose deviation is not necessarily effective in predicting the need for re-planning for all patient populations. Lastly, the high fidelity delivered dose data presented here should be useful for exploring novel strategies to most effectively predict the need for and timing of re-planning efforts; a topic of future work for our group.

## 5. Conclusions

Our use of gold standard FBCT image data allowed for characterization of the total, delivered dose for each of the 74 HNC patients studied here with accuracy comparable to the original simulation-based dose calculation and, thereby, eliminated the uncertainties of previous CBCT-based studies. The accumulated, total delivered-dose distributions agreed well for the vast majority of patients in this dataset. However, clinically notable deviations were observed for the summed delivered dose to the parotid glands of ten patients, leading to NTCP increases of 11% − 44%. We further determined that a mid-course evaluation of dose deviation was not effective in predicting the need of re-planning for our patient cohort.

The high fidelity, FBCT-based dose data presented here should be extremely useful for exploring novel strategies to most effectively predict the need for and timing of re-planning efforts; a topic of future work for our group.

Therefore, it is important to appreciate how inherent and unavoidable setup discrepancies, combined with anatomical changes over time, can manifest as non-trivial deviations of the intended, delivered dose. These non-trivial increases to parotid gland delivered dose suggest that even when rigorous, high quality image guidance is performed, clinically concerning variations to predicted dose delivery can still occur.

## Data Availability

All data produced in the present study are available upon reasonable request to the authors

## Notes

### Competing Interest Statement

The authors have declared no competing interest.

### Funding Statement

This study did not receive any funding

### Author Declarations

IRB of the University of Utah gave ethical approval for this work

